# Exploration the Standard diagnosis and treatment in Patients with Type 2 Diabetes in a tertiary first-class hospital in Fujian Province

**DOI:** 10.1101/2020.03.06.20026591

**Authors:** Xiaoyan Lin, Wei Lin, Junping Wen, Gang Chen

## Abstract

**Objective:** The aim of this study was to evaluate the knowledge and clinical practice of ‘‘guideline for prevention and treatment of Type 2 diabetes in China’’ among the physicians. We took a tertiary first-class hospital in Fujian Province as an example to identify the differences between clinical practice and guideline.

**Methods:** We selected 3000 inpatients with type 2 diabetes who visited at the tertiary first-class hospital in Fujian Province between September 2017 to November 2017, then retrospectively analyzed their clinical data, including age, gender, height, weight, body mess index, admission departments, combined diseases, complications, course of disease, diabetes diet education, monitoring of blood glucose and blood pressure, the selection of hypoglycemic therapy and other secondary prevention measures. The data we obtained was analyzed by SPSS 19.0 software.

**Results:** A total of 3000 effective inpatient were enrolled, including 1724 male and 1276 female, the mean age(± standard deviation) of patients was 66.21 ± 11.75 years, the mean hospitalization days was 12.26±10.30 days.In this study, we found that:1.only 60.9% of the patients have monitored HbA1C in nearly 3 months; in the last year, 96.3% of patients have monitored blood lipid;98.1% of patients have monitored the serum creatinine;only 9.5% of patients have received comprehensive eye examination by an ophthalmologist or optometrist.2.After admission, 73.3% of patients have received the education about diabetes diet; 86.8% of patients have accepted the peripheral blood glucose monitoring;among 2084 diabetic patients combined with hypertension, only 1868 patients (89.6%) have received blood pressure monitoring.3. Among 202 patients who are newly diagnosed diabetes, there were 45 cases meet HbA1C > 9.0% or FPG > 11.1 mmol/ L, only 25 people (55.56%) have received an intensive insulin therapy.4.Among 474 patients who have undergone an large or medium-sized surgical treatment, about 182(62.1%) patients have started a hypoglycemic therapy before the operation, and only 196(41.4%) patients have changed into intensive insulin therapy before surgery.5. About 714 of diabetic patients who combined with a clear history of cardiovascular, only 548 (76.8%) patients have received antiplatelet therapy;and 566 people (79.3%) have used statin to lower blood lipid. 6.In total, only 168(5.6%) patients have received a relatively standard diagnosis and treatment in non-surgical department, and only 4(0.1%) patients in surgical department.

**Conclusion:** There are still many deficiencies in clinical practice of type 2 diabetes guideline, even in tertiary first-class hospital, there are also many physicians, especially non-specialist doctors, were not familiar with the knowledge of guideline for management of Type 2 diabetes.The acute exacerbation of diabetes in the global can take serious complications and bring heavy economic burden, so it required us to pay attention to the prevention and early treatment of diabetes. We suggest to popularize the “guideline for management of Type 2 diabetes” through various forms, such as academic seminars, lecture tour and medical staffs, so as to enhance the education of physicians and promote treatment based on guideline of type 2 diabetes.

## Introduction

Diabetes mellitus has already become a serious world-wide public health problem, due to the large number of patients with diabetes, the following complicated complications, and the heavy socio-economic burden.In recent years, diabetes began to spread aroud the world, because of the extension of life expectancy, the improvement of living standards, the decrease of physical activity and the transformation of the life-style^[1-2].^ According to statistics, the number of diabetics in the world was 120 million in 1994, 137 million in 1997 and 175 million in 2000. It is predicted that the number of people suffering from diabetes in 2030 will reach 300 million^[3]^.As China is a country with a large population in the world, its diabetic population is not to be underestimated. According to a recent study, about 92.4 million adults over the age of 20 in China have diabetes (equivalent to 9.7 percent of adults), of which 60.7 % have not yet been diagnosed.With the increase in the prevalence of diabetes, the economic burden is getting heavier and heavier. According to the WHO (World Health Organization, World Health Organization), by 2025, the medical cost of diabetes will account for as 13% of the global health budget, and will account for 40% in countries with a high incidence of diabetes^[4]^.Another domestic study showed that the total medical cost of diabetes in China increased from 2.216 billion yuan in 1993 to 200 billion yuan in 2007, and the proportion of total health expenditure also increased from 1.96 to 18.2 percent, an eight-fold increase in just 15 years^[5]^.

In order to curb the prevalence of diabetes, to reduce the incidence and mortality of it’s chronic complications, so as to reduce the socio-economic burden.Since 1991, experts in relevant fields at home and abroad have been actively organized to draw up a guideline to consistent with different national conditions or various countries, which based on the results of large-scale randomized controlled clinical trials, in order to provide guidance and suggestions for clinical medical workers.The same is true in China. Since 2003, the CDS (Chinese Diabetes Society) has organized national experts to draw lessons from the latest research results and guiding suggestions at home and abroad, and combined with the recent epidemiological and syndrome medical results in China. The guidelines for the Prevention and treatment of Type 2 Diabetes in China were compiled and updated in 2007, 2010, 2013 and 2017, respectively.

Although the blood sugar has been greatly improved in the following years compared with before the introduction of the guidelines, the overall control situation is still not satisfactory^[5]^. A cross-sectional survey in the United States followed 4926 diabetics over the age of 20 and found that between 2007 and 2010, only 56.2 per cent of patients achieved the target of less than 7.0 percent of HbA1c. The blood pressure was controlled below 130/80mmHg in 51.1% of the patients, LDL < 100mg / dL in 56.2% of the patients, and only 18.8% of the patients reached the three standards at the same time^[6]^.At present, the China guidelines for the prevention and treatment of Type 2 Diabetes (2013 Edition) has been in force for more than five years, but no relevant research has been conducted to investigate its clinical implementation. This paper reviews the relevant clinical data of inpatients with type 2 diabetes mellitus in a tertiry A hospital in Fujian Province from June 26, 2017 to November 06, 2017. To find out whether all inpatients with type 2 diabetes mellitus received individualized treatment under the guidance of guidelines, and then reflect the mastery and implementation of the guidelines by all doctors in our hospital.

## Data and methods

### 1. Research object

By using the method of cluster sampling, all patients with type 2 diabetes who were admitted to the inpatient department of a tertiry A hospital in Fujian Province from June 26, 2017 to November 06, 2017 were selected, including patients with type 2 diabetes who were previously diagnosed or newly diagnosed after admission. Patients who were discharged from the hospital for only one day or were discharged from the day ward, who were suspected of diabetes and diagnosed as other types or diagnosed as unkown type were excluded.The diagnostic criteria of diabetes for newly admitted patients refer to the ‘‘guideline for prevention and treatment of Type 2 diabetes in China (2013 Edition)’’(The following is called a guide):

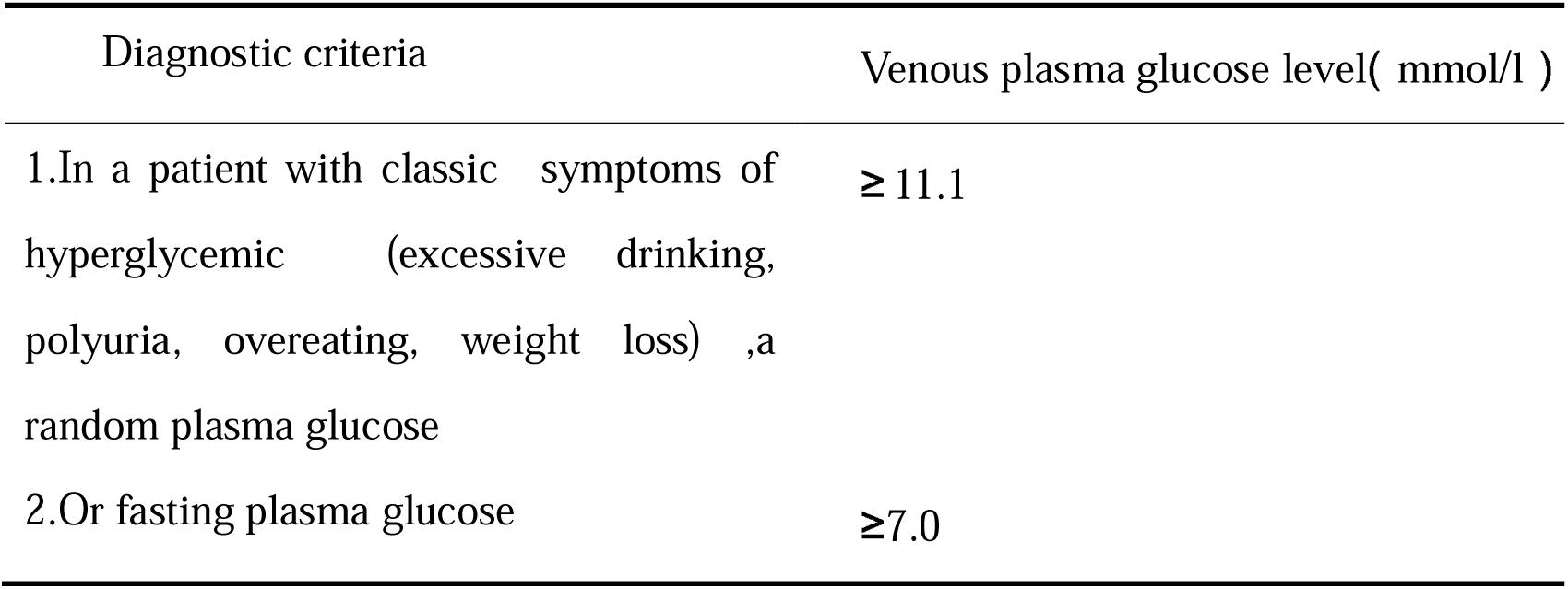
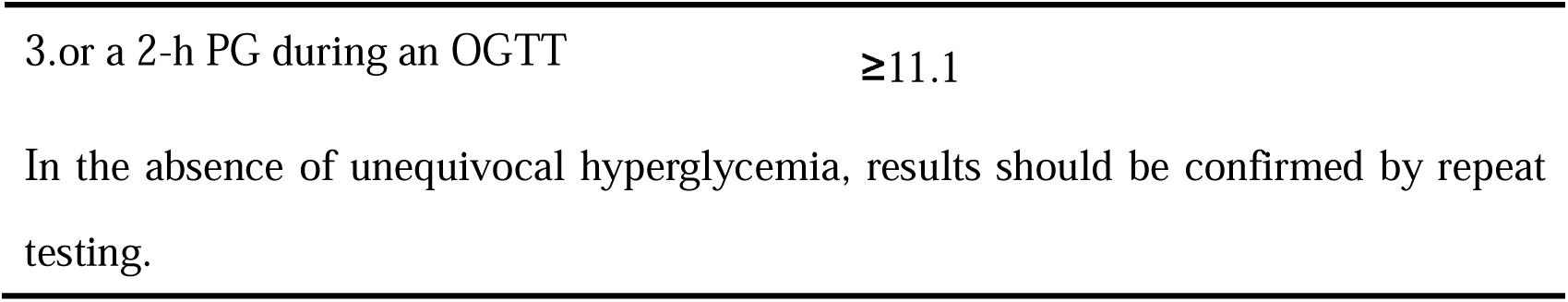

The diagnostic criteria of hypertension refer to the “guidelines for the prevention and treatment of hypertension in China (2010 Edition)” ^[7]^: without using of antihypertensive drugs, systolic blood pressure ≥ 140mmHg and / or diastolic blood pressure ≥ 90mmHg which were measured three times on the different day. Blood pressure was measured in supine position, using standard mercury pump.BMI (Body mass index)= weight (kg) / {height × height (m2)}. The biochemical indexes such as fasting blood glucose, triglyceride, total cholesterol, low density lipoprotein, urea nitrogen and creatinine were determined by automatic analyzer using Chemiluminescence method. All biochemical indexes were tested in the next morning when patient were fasting.Fasting is defifined as no caloric intake for at least 8 hours. In the end, a total of 3000 patients with type 2 diabetes were diagnosed as “type 2 diabetes”, with or without other diseases, including 1724 males and 1276 females with an average age of (66.21 ±11.75) years. The average hospital stay was (12.26 ±10.30) days, the average height was (163.47 ±7.93) cm, the average weight was (64.26 ±10.26) Kg, the average BMI was (24.04 ±4.85) Kg/m2 (more basic information see Table 1).

**Table 1.**
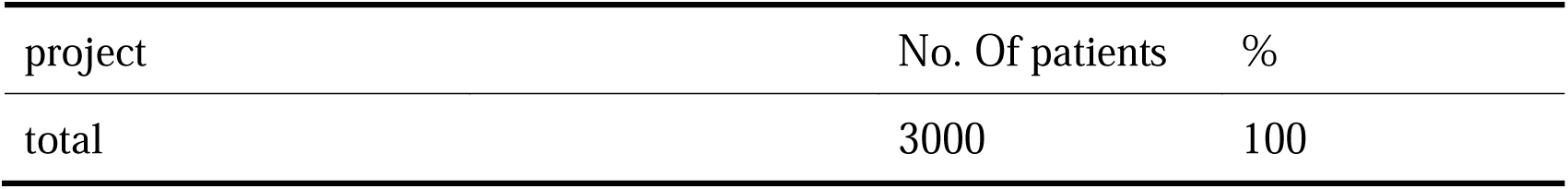
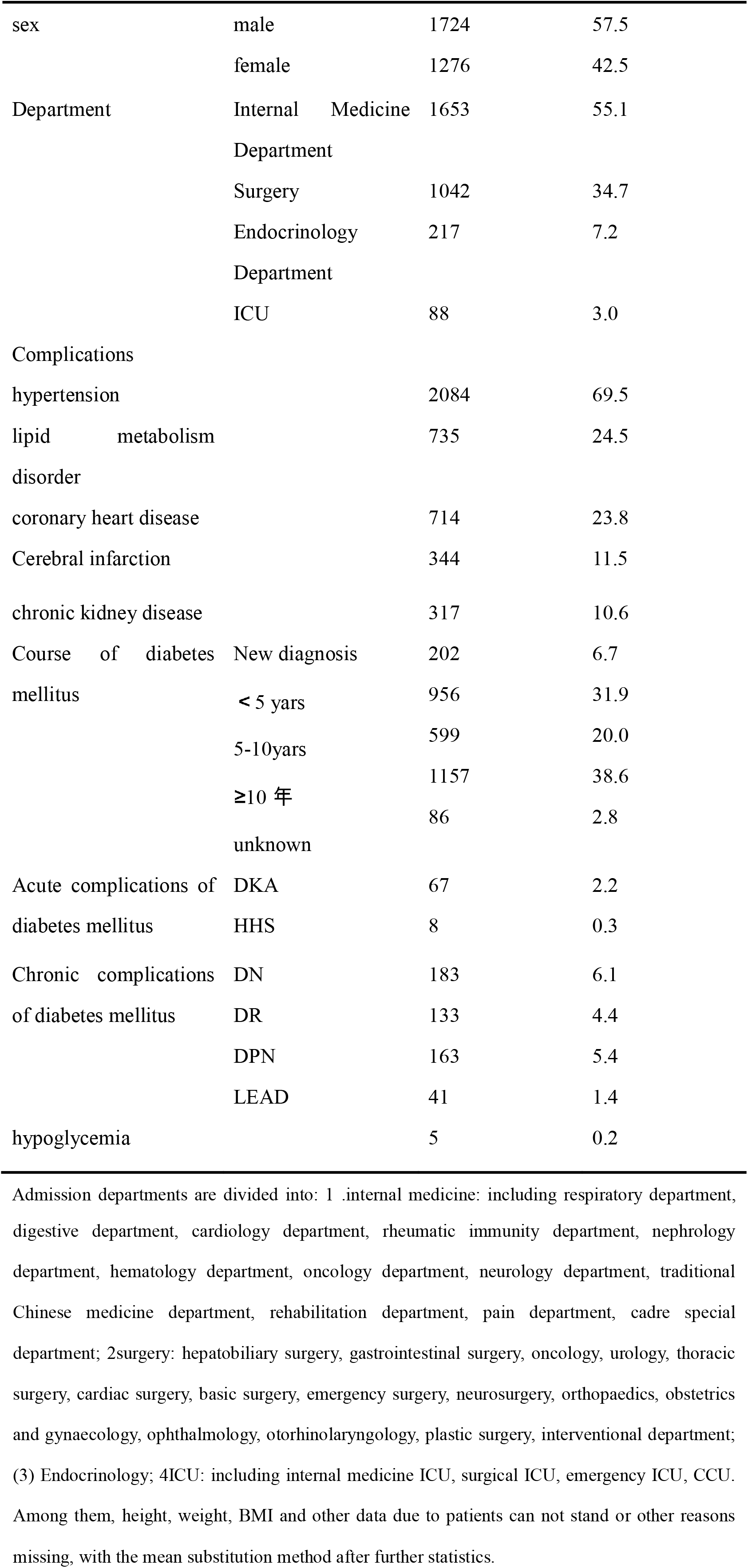
Patient basic information

**Table 1.1.**
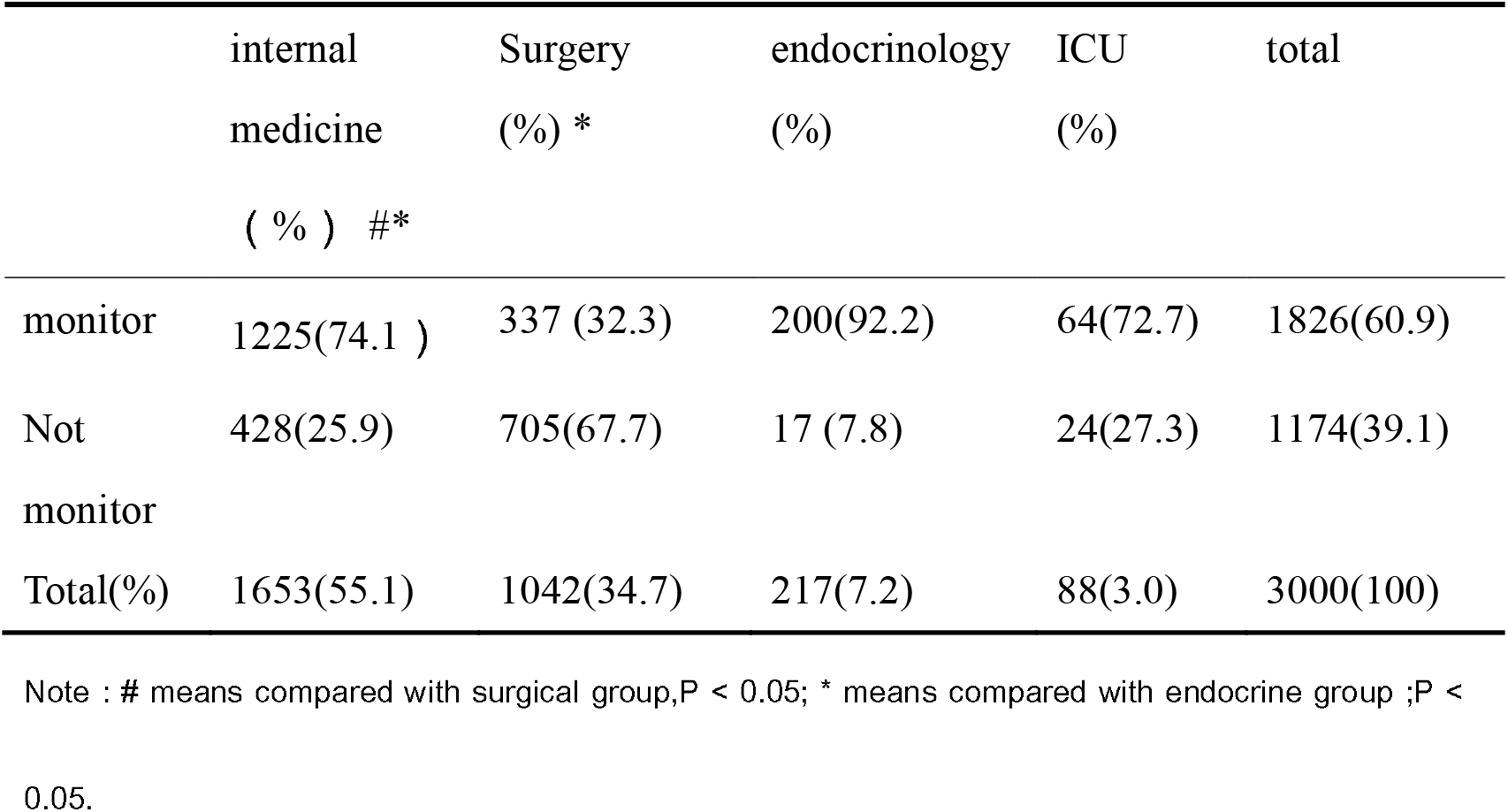
HbA1C monitoring of the target population in recent 3 months

**Table 1.2.** Monitoring of blood lipids in the target population

|  | internal<br>medicine<br>( % ) | Surgery<br>( % ) | endocrinology<br>(% ) | ICU<br>( % ) | Total<br>( % ) |
| --- | --- | --- | --- | --- | --- |
| monitor | 1610(97.4) | 981(94.2) | 211(97.2) | 86(97.7) | 2888(96.3) |
| Not<br>monitor | 43(2.6) | 61(5.8) | 6(2.8) | 2(2.3) | 112(3.7) |
| total ( % ) | 1653(55.1) | 1042(34.7) | 217(7.2) | 88(3.0) | 3000(100) |

Note:There was no significant statistical significance in the monitoring of blood lipids in each group.

**Table 1.3.**
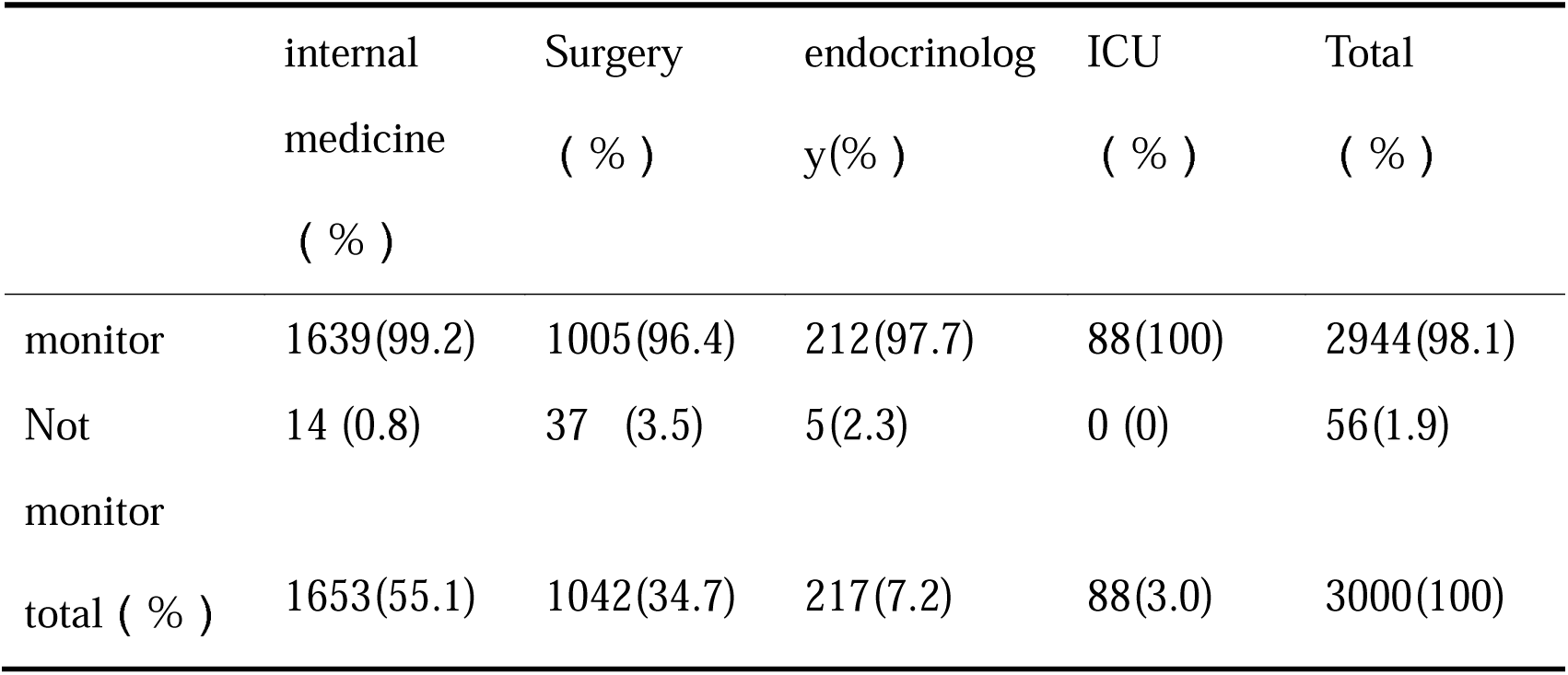
Monitoring of serum creatinine in target population

**Table 1.4.** Screening of urinary Microalbumin in the past one year in the included population

|  | internal<br>medicine<br>( % ) #* | Surgery<br>( % ) * | endocrinolog<br>y( % ) | ICU<br>( % ) | Total<br>( % ) |
| --- | --- | --- | --- | --- | --- |
| monitor | 179(10.8) | 17(1.6) | 144(66.4) | 4(4.5) | 344(11.5) |
| Not<br>monitor | 1474(89.2) | 1025(98.4) | 73(33.6) | 84(95.5) | 2656(88.5) |
| total ( % ) | 1653(55.1) | 1042(34.7) | 217(7.2) | 88(3.0) | 3000(100) |

Note : # means compared with surgical group,P < 0.05; \* means compared with endocrine group ;P < 0.05.

**Table 1.5.** Fundus evaluation of included population

|  | internal<br>medicine<br>( % ) #* | Surgery<br>( % ) * | endocrinolog<br>y( % ) | ICU<br>( % ) | Total<br>( % ) |
| --- | --- | --- | --- | --- | --- |
| Evaluation | 106 ( 6.4) | 36(3.5) | 145(66.8) | 0(0) | 287(9.5) |
| Not<br>Evaluation | 1549(93.7) | 1006(96.5) | 72(33.18) | 88(100) | 2715(90.5) |
| Total ( % ) | 1653(55.1) | 1042(34.7) | 217(7.2) | 88(3.0) | 3000(100) |

Note : # means compared with surgical group, $P < 0.05$ ; \* means compared with endocrine group ; $P < 0.05$ .

## 2. Methods

By using the method of retrospective analysis, we try to understand the knowledge, attitude and implementation of diabetes guidelines among clinicians in the departments of tertiary first-class hospital through the comprehensive analysis and statistics of the general data, past medical history, laboratory examination, nursing records and medical orders of the included population. This study is approved by IRB of Fujian Provincial Hospital.

## 3. Statistic analysis

All the data were imported into Excel 2010, corrected and re-recorded after input, and imported into SPSS 19.0 software for statistical analysis. The counting data were described by constituent ratio (%), the measurement data were statistically described by x ±s.The measurement data were measured by t-test, and the counting data were measured by Chi-square test (P < 0.05). There was significant difference between the two groups (P < 0.05).

## Result

### 1. The evaluation of disease on admission

#### 1.1 Monitoring of glycosylated hemoglobin

According to the guideline, HbA1C (Glycosylated Hemoglobin) should be monitored every 3 months at the initial stage of diabetes treatment, before stable blood glucose control, or during the replacement of antihyperglycemic regimens; and at least twice a year after blood glucose is up to standard. By analysis the previous history and laboratory examination after admission, we finded that only 60.9% (1828) of the patients had been monitored HbA1C in past 3 months.In the rest of 39.1% (1172) patients who were not monitored HbA1C, internal medicine department accounted for 14.3%, surgery department accounted for 23.5%, endocrinology department accounted for 0.5%, and ICU accounted for 0.8% (see Table 1.1).

#### 1.2 Monitoring of Blood Lipid

The guideline recommend that all diabetics should monitor blood lipids (including total cholesterol, low density lipoprotein cholesterol, and triglycerides) at least once a year. According to the statistics, 96.4% (2888) of the target patients have received blood lipid examination in the past one year, only 3.6% (112) of the patients didn’t check their blood lipid levels (see Table 1.2).

**Table 2.1.**
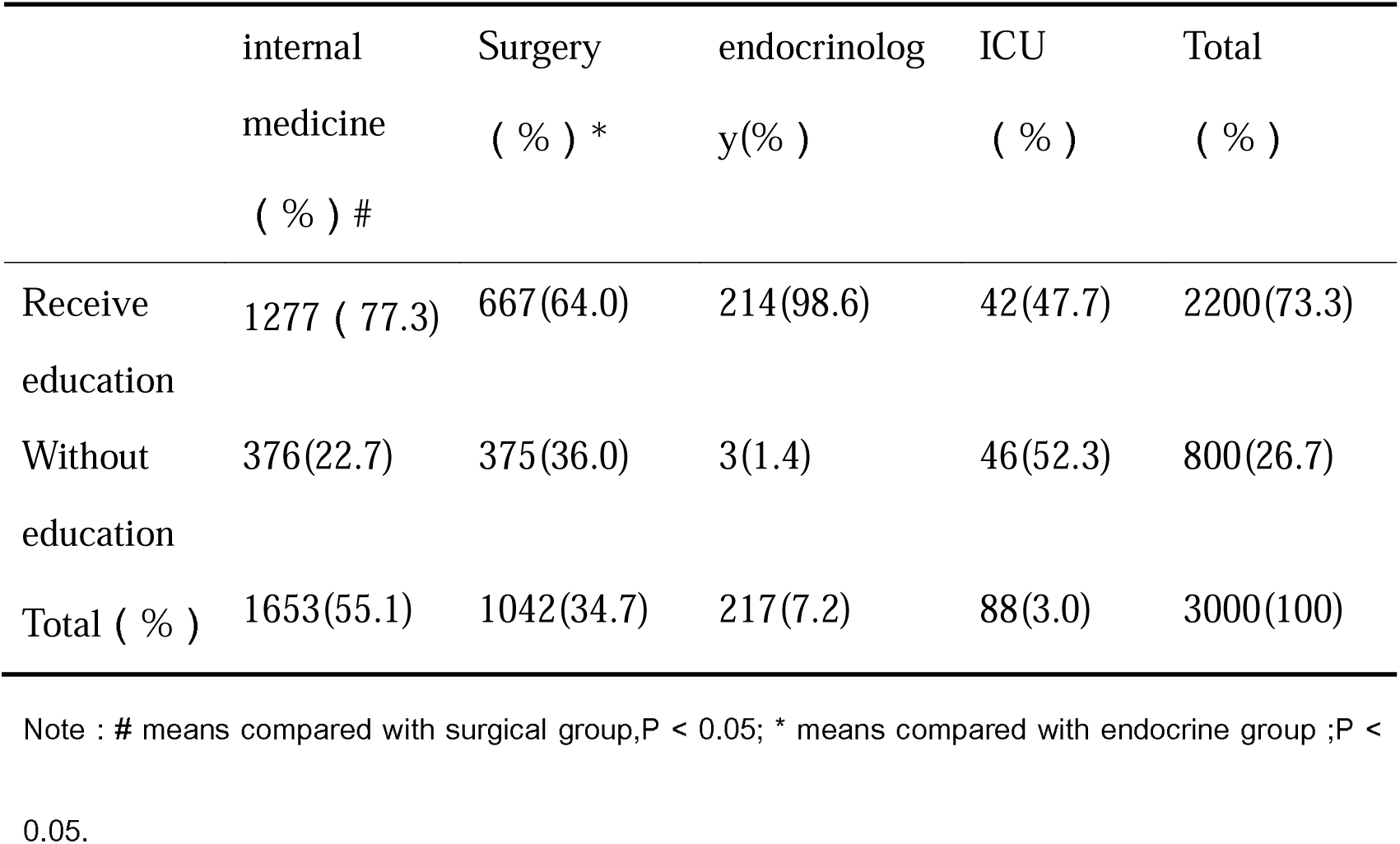
diabetes diet education in the population

|  | internal<br>medicine<br>( % ) # | Surgery<br>( % ) * | endocrinolog<br>y( % ) | ICU<br>( % ) | Total<br>( % ) |
| --- | --- | --- | --- | --- | --- |
| Receive<br>education | 1277 ( 77.3) | 667(64.0) | 214(98.6) | 42(47.7) | 2200(73.3) |
| Without<br>education | 376(22.7) | 375(36.0) | 3(1.4) | 46(52.3) | 800(26.7) |
| Total ( % ) | 1653(55.1) | 1042(34.7) | 217(7.2) | 88(3.0) | 3000(100) |

Note : # means compared with surgical group, $P < 0.05$ ; \* means compared with endocrine group ; $P < 0.05$ .

**Table 2.2.**
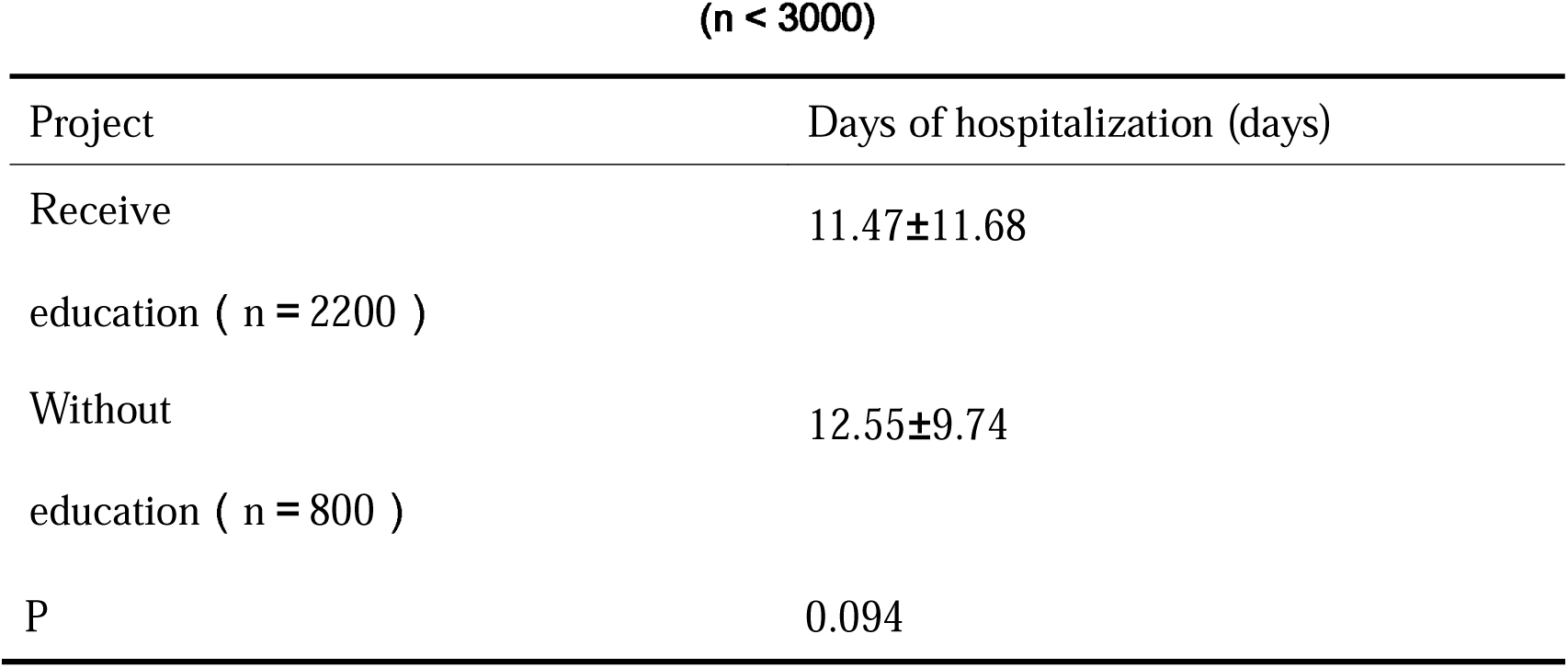
comparison of hospitalization days of patients with or without dietary education (n < 3000)

**Table 2.3.** inclusion of blood glucose monitoring in the population

|  | internal<br>medicine<br>( % ) # | Surgery<br>( % ) * | endocrinolog<br>y( % ) | ICU<br>( % ) | Total<br>( % ) |
| --- | --- | --- | --- | --- | --- |
| monitor | 1464 ( 88.6 ) | 838 ( 80.4 ) | 215 ( 99.1 ) | 86 ( 97.7 ) | 2603 ( 86.8 ) |
| Not<br>monitor | 189 ( 11.4 ) | 204 ( 19.6 ) | 2 ( 0.9 ) | 2 ( 2.3 ) | 397 ( 13.2 ) |
| total ( % ) | 1653 ( 55.1 ) | 1042 ( 34.7 ) | 217 ( 7.2 ) | 88 ( 3.0 ) | 3000 ( 100 ) |

Note : # means compared with surgical group, $P < 0.05$ ; \* means compared with endocrine group ; $P < 0.05$ .

**Table 2.4.** comparison of hospitalization days in patients with or without blood glucose monitoring (n < 3000)

| Project | Days of hospitalization (days) |
| --- | --- |
| monitor ( n = 2603 ) | 8.77±10.02 |
| Not monitor ( n = 397 ) | 12.79±10.25 |
| P | 0.015 |

**Table 2.5.** Blood pressure monitoring in the target population

|  | Monitor blood<br>pressure ( % ) | Not Monitor blood<br>pressure ( % ) | Total ( % ) |
| --- | --- | --- | --- |
| Complicated with<br>hypertension | 1868(80.0 ) | 216(32.5 ) | 2084(69.5) |
| Not complicated with<br>hypertension | 467(20.0) | 449(67.5 ) | 916(30.5) |
| Total ( % ) | 2335 ( 77.8 ) | 665 ( 22.2 ) | 3000(100 ) |

**Table 2.6.**
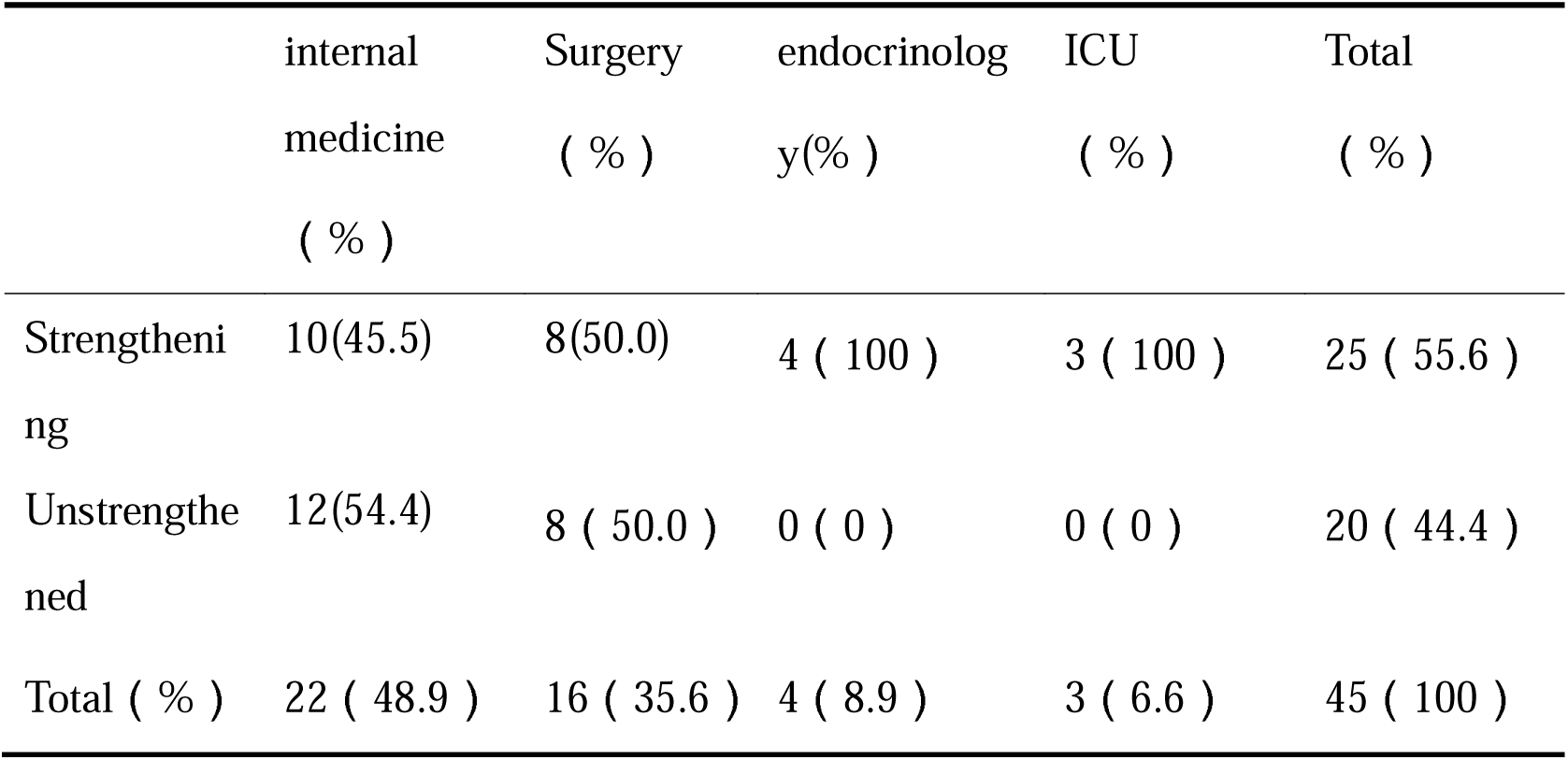
HbA1C > 9.0% or FPG > 11.1mmol/L in newly diagnosed diabetic patients with intensive hypoglycemic therapy

**Table 2.7.**
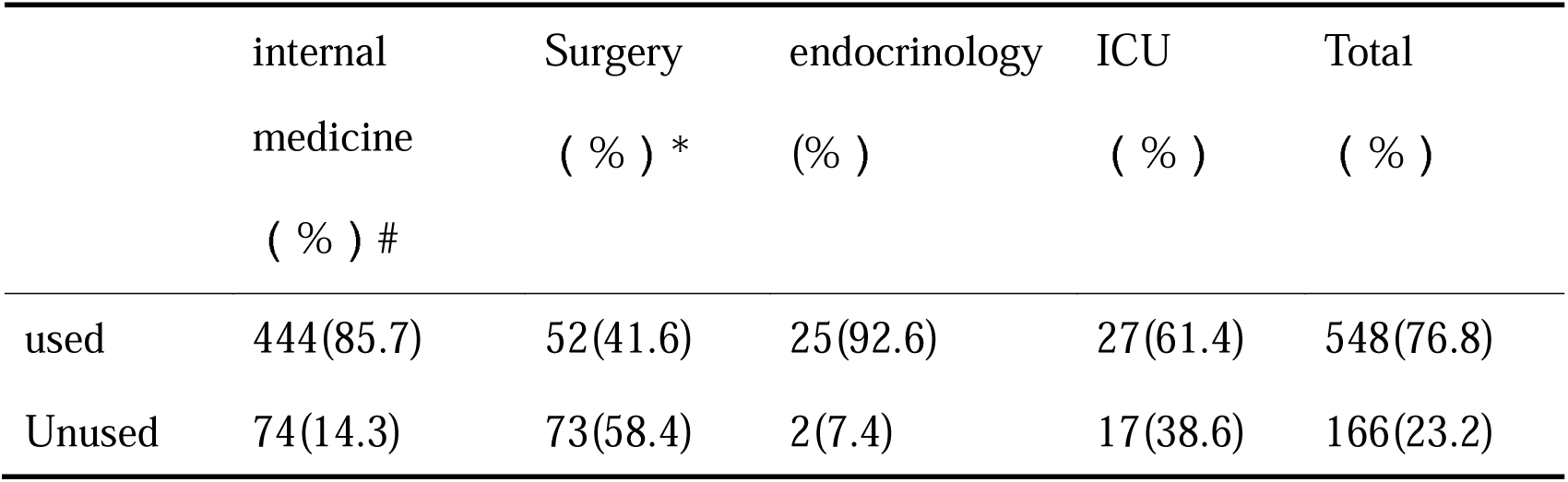
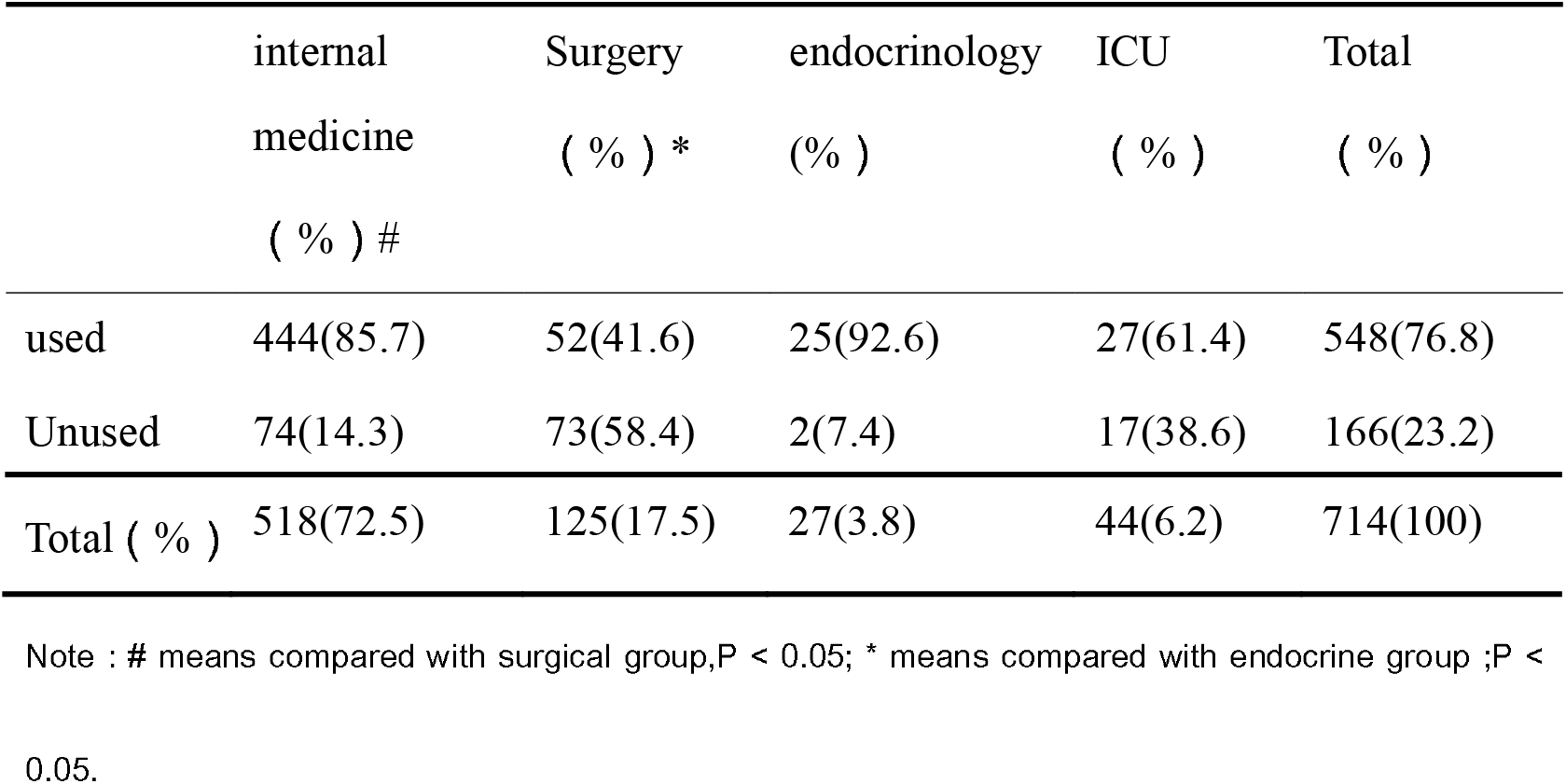
The use of antiplatelet drugs in diabetic patients with a history of cardiovascular disease

**Table 2.8.** The use of statins in diabetic patients with a history of cardiovascular disease

|  | internal<br>medicine<br>( % ) # | Surgery<br>( % ) * | Endocrinology<br>(% ) | ICU<br>( % ) | Total<br>( % ) |
| --- | --- | --- | --- | --- | --- |
| used | 443(85.5) | 68(54.4) | 24(88.9) | 31(70.5) | 566(79.3) |
| Unused | 75(14.5) | 57(45.6) | 3(11.1) | 13(29.5) | 148(20.7) |
| Total ( % ) | 518(72.5) | 125(17.5) | 27(3.8) | 44(6.2) | 714(100) |

Note : # means compared with surgical group,P < 0.05; \* means compared with endocrine group ;P < 0.05.

#### 1.3 Determination of serum creatinine

According to the guidelines, regardless of the level of urinary protein excretion, all adult diabetic patients need to test for creatinine at least once a year, to calculate eGFR (Estimated glomerular filtration Rate), so as to assess whether they are complicated with chronic kidney disease. In the target population, only 1.9% (56) patients did not monitor serum creatinine (see Table 1.3).

Urinary microalbumin is a more sensitive indicator for the detection of diabetic, and further statistics were carried out in the target population.,It was found that only 11.5% (344) of patients had tested urinary microalbumin in the past year (see table 1.4).

#### 1.4 Evaluation of diabetic retinopathy

DR (Diabetes retinopathy) is a specific microangiopathy of diabetic. According to statistics, only 9.5% (287) patients have recieved a consultation and the fundus examination by professional ophthalmologists. Among the 90.5% (2715) of the patients who didn’t receive fundus examination, 51.6% were in internal medicine department, 33.5% in surgery department, 2.4% in endocrine department and 3.0% in ICU (see table 1.4).

**Table 4.1.**
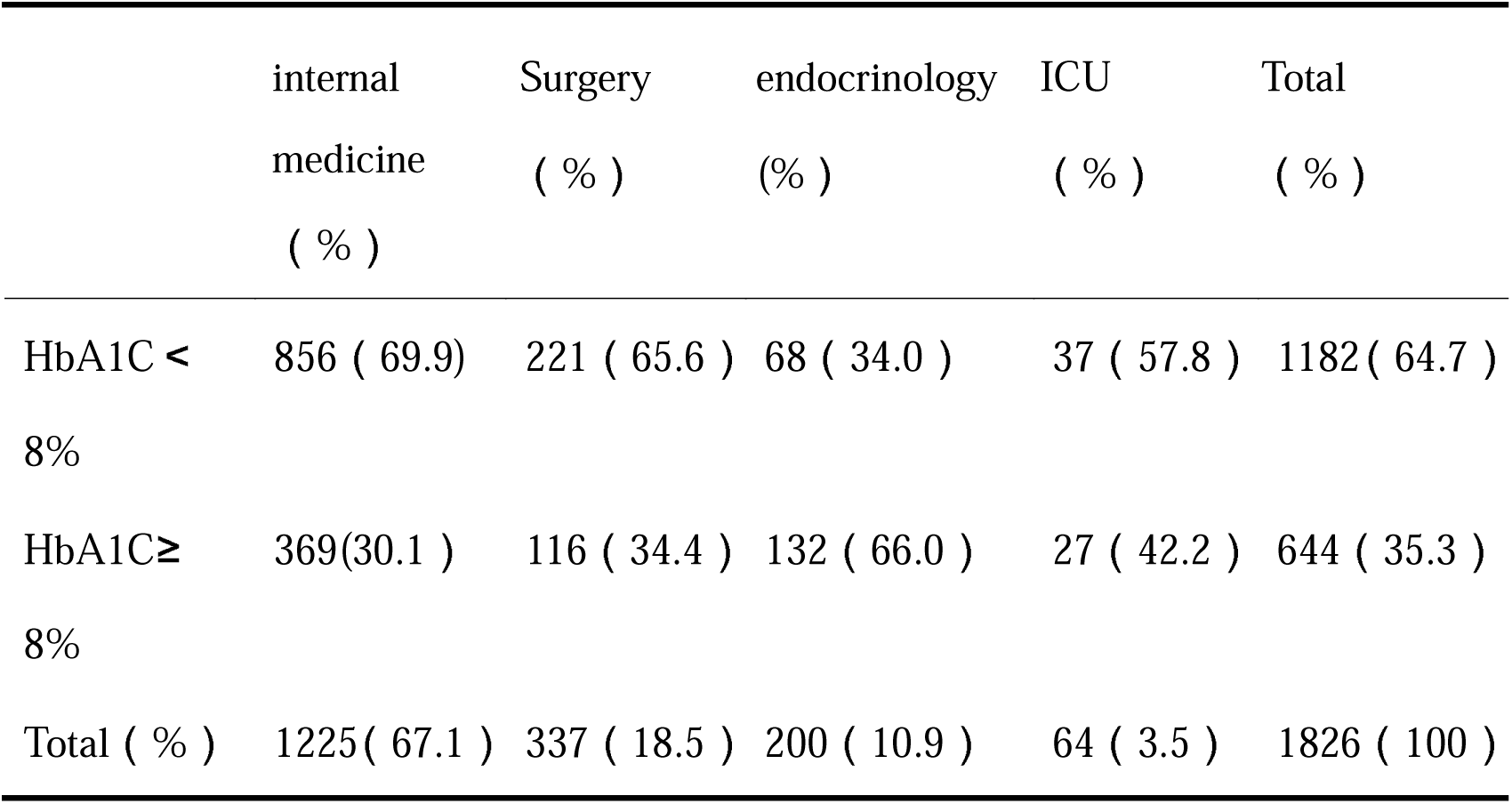
HbA1C status of the included population at admission

**Table 4.2.** Blood glucose compliance of diabetic patients before discharge

|  | internal<br>medicine<br>( % ) # | Surgery<br>( % ) * | Endocrinolo<br>gy<br>(% ) | ICU<br>( % ) | Total<br>( % ) |
| --- | --- | --- | --- | --- | --- |
| standards | 555( 50.6 ) | 415 ( 54.7 ) | 123(62.8) | 10(23.3) | 1103(52.6 ) |
| substandard | 542 ( 49.4 ) | 344 ( 45.3 ) | 73(37.2) | 33(76.7) | 992 ( 43.4 ) |
| Total ( % ) | 1097 | 759 | 196 | 43 | 2095(100) |

Note : # means compared with surgical group, P < 0.05; \* means compared with endocrine group ; P < 0.05.

**Table 4.3.** Blood pressure control in diabetic patients with hypertension before discharge

|  | internal<br>medicine<br>( % ) | Surgery<br>( % ) | Endocrinology<br>( % ) | ICU<br>( % ) | Total<br>( % ) |
| --- | --- | --- | --- | --- | --- |
| standards | 634(65.6) | 289(63.8) | 87(68.0) | 34(65.4) | 1044(65.3) |
| substandard | 332(34.4) | 164 (36.2) | 41(32.0) | 18(34.6) | 555(34.7) |
| Total ( % ) | 966(60.4) | 453(28.3) | 128(8.0) | 52(3.3) | 1599(100) |

### 2. Comprehensive treatment of diabetes mellitus

#### 2.1 Diabetes diet education

As the basic treatment of diabetes, diet therapy is essential for the smooth control of blood glucose. However, in real life, there are very few patients who can adhere to diet therapy. In 2010^[8]^, a survey from Changchun City Jilin Province showed that among 270 community diabetic patients, 24 (8.89%) patients had good dietary compliance and 49 (18.11%) patients had moderate compliance, the rest of 197 cases (73%) had poor compliance.We can see that the overall diet control of diabetic patients is not optimistic. So, how many clinicians will give dietary guidance and education to patients after admission? In this hospital, the nurse will carry out the diet guidance and education to the patient after the doctor gives the diet order, Therefore, we collected the doctor’s order, and found that 73.4% (2200 cases) of the population had been given the order for diabetes diet.In the rest of 26.7% (800cases) who without diet education, 12.5% were in internal medicine department and 12.5% in surgery department, and endocrine department accounted for 0.1%, ICU accounted for 1.5% (see Table 2.1).

There is no significant difference in the days of hospitalization between the two groups.

#### 2.2 Blood glucose monitoring

In order to make diabetic patients adjust their blood glucose more conveniently and avoid the attack of hypoglycemia, dynamic monitoring of blood glucose is an essential measure. Several studies have confirmed the role of SMBG (Self-monitoring of blood glucose, self-monitoring of blood glucose) in blood glucose control in patients with type 2 diabetes ^[9-10]^. In this survey, regardless of frequency and time of blood glucose monitoring weather correct or not, a total of 86.7% (2603) of patients have received peripheral blood glucose monitoring after admission. The statistics of 13.2% (397) of the patients who did not receive blood glucose monitoring were as follows: internal medicine department account for 6.3%, surgery department account for 6.8%, endocrinology account for 0.1%, ICU 0.1% (see Table 2.3).

#### 2.3 Blood pressure monitoring

We conducted statistics on blood pressure monitoring in the included population as follows (see table 2.5). Further analysis on the rest of 10.5% (216/2084) hypertension patients without blood pressure monitoring showed that internal medicine department accounted for 5.9%, surgery department 4.4%, endocrine department accounted for 0.1%, ICU accounted for 0.1%.

#### 2.4 Are newly diagnosed diabetic patients with HbA1C > 9.0% or FPG > 11.1mmol/L treated with insulin for intensive hypoglycemic therapy?

There are many kinds of antihyperglycemic drugs and a variety of hypoglycemic schemes, so we take the newly diagnosed diabetes as an example for analysis. According to the guidelines, short-term intensive insulin therapy is recomended for newly diagnosed type 2 diabetic patients with HbA1C > 9.0% or FPG > 11.1mmol/L. A total of 202 newly diagnosed diabetic patients were included, of which 45 met HbA1C > 9.0% or FPG > 11.1 mmol / L, the statistics of whether or not to adopt intensive hypoglycemic regimen in their departments are as follows (see Table 2.6).

#### 2.5 The management of blood glucose in Peri-operative

We defined the surgery which duration is more than 1 hour or use the intraspinal anesthesia or general anesthesia or required fasting before surgery as large or medium-sized surgery, about 474 patients were included. According to the guideline, patients who are suffer from the major or moderate surgery should stop taking oral antihyperglycemic drugs three days before operation and change to insulin therapy. Therefore, we analyzed the preoperative antihyperglycemic regimen in the population and found that in 474 included patients, 61.8% (293/474)patients had started antihyperglycemic therapy before operation, including oral hypoglycemic drugs and subcutaneous insulin injection, and only 62.1% (182-293) of patients were adjusted to insulin therapy to reduce blood glucose (including basal insulin, continuous insulin pump or basal + meal insulin or premixed insulin multiple times a day).

Because most of the inpatients are elective surgery, it is generally recommended to control the preoperative blood glucose below 11.1mmol/L. Four consecutive times of blood glucose ≥ 3 times below 11.1mmol/L were defined as the standard of blood glucose before operation. In the 474 included patients in this study, only 41.4% (196/474) patients were reach to the standard of blood glucose.In the same, four consecutive times of postoperative blood glucose ≥ 3 times below 10.0mmol/L were confined as achieve blood glucose standard. After analysis, there are only 51.7% (245/474) of the patients reached the postoperative blood glucose standard. We can see that there are also about half of the surgeons didn’t have a good grasp of the guidelines and can’t implement them well, no matter in the adjustment of antihyperglycemic regimen or the control of perioperative blood glucose.

#### 2.6 Secondary prevention of diabetes mellitus with a history of cardiovascular disease

Due to the hypercoagulable state of diabetic and the disorder of glucose and lipid metabolism, cardiovascular disease is a common macrovascular Complications in diabetic patients. It has been proved by many trials and meta analysis that aspirin has been gradually recognized as a secondary prevention of diabetes mellitus complicated with cardiovascular disease. In the 714 patients with a clear history of cardiovascular disease, only 76.8% (548) patients had received antiplatelet therapy (see Table 2.7 for details). Further analysis of 23.2% (166) patients who did not receive secondary prophylaxis with antiplatelet drugs showed that 1.3% (10 patients) patients were complicated with gastrointestinal bleeding and 0.6% (4 patients) patients complicated with digestive system ulcer, 4.8% (34 cases)patients stop using drugs becouse of operation, 0.4% (3 cases) patients complicated with Cerebral hemorrhage, and the rest of 16.1% (115 cases) patients without Significant contraindication.

#### 2.7 The use of statins in diabetic patients with a history of cardiovascular disease

The management of blood lipids is also an important factor which can affect the recurrence and prognosis of cardiovascular disease. The guideline recommend that diabetic patients with a clear cardiovascular disease should use statins on the basis of lifestyle intervention. According to the statistics of the included population, there are 714 diabetic patients with cardiovascular disease, only 79.3% (566) patients were treated with statins (see Table 2.8 for details).

## 3. The implementation rate of the guideline

Suppose there are following conditions: □Blood lipids and serum creatinine were monitored in the past 1 year, fundus was evaluated in the past 1 year, glycosylated hemoglobin was monitored in the past 3 months;□patients were received a diet education after admission; ➂peripheral blood glucose and blood pressure were monitored after admission.□ patients with coronary heart disease were treated with statins; □ patients with coronary heart disease were treated with antiplatelet therapy; ➅ patients with newly diagnosed diabetes mellitus with HbA1C > 9.0% or FPG > 11.1 mmol / L were treated with intensive hypoglycemic therapy;➆the perioperative patients had adjusted their antihyperglycemic scheme, and the blood glucose was controlled well before and after operation.If the surgical inpatients who met the conditions of 1, 2, 3, 4, 6, 7 at the same time, we defined it as good execution of the guidelines. While, in non-surgical patients who meet the conditions of 1, 2, 3, 4, 5, 6 at the same time are defined as good implementation of the guidelines. Our analysis showed that only 5.6% (168/3000) of the non-surgical patients had implemented the guideline well, including 1.9% (58-3000) in internal medicine department and 3.7% (110-3000) in endocrine department. All the patients admitted in ICU were unqualified because of no fundus examination. Only 0.1% (4/3000) of the surgical patients had implemented the guideline well, and all the patients who underwent surgery did not meet the requirements because of the poor control of perioperative blood glucose.

## 4. The control of diabetes mellitus among inpatients

### 4.1 Glycosylated hemoglobin on admission

Because the average age of the included population was (66.21 ±11.75) years, most of them were not newly diagnosed diabetes mellitus, the course of disease was longer, and they were often complicated with cardiovascular and cerebrovascular diseases, tumors or other complex basic diseases. Therefore, we use the relatively loose HbA1c target (HbA1C < 8%) to define whether HbA1c is well controlled or not. We statistically analyzed the 1828 people who had monitored for HbA1c after admission as follows (see Table 3.1):

### 4.2 Blood glucose control at discharge

The blood glucose level at the time of discharge can be used to evaluate the effect of antihyperglycemic therapy during the whole period of hospitalization. We counted the last four times of peripheral blood glucose continuously monitored before discharge. We defined that more than 3 times of peripheral blood glucose control within the range of 3.9-10mmol/l as blood glucose control standard, otherwise, the blood glucose control was not up to the standard. We reviewed the blood glucose level of the included population monitoring, and after excluding the unmonitored, unrecorded population for various reasons, a total of 2095 people were included, as follows (see Table 3.2 for details):

### 4.3 Blood pressure control at discharge

Similarly, the control of blood pressure is also important for the prognosis of diabetic patients and the prevention of cardiovascular complications. We analyzed the blood pressure of the last 4 times of continuous monitoring before discharge. We defined the patient as standard, whose blood pressure are below 140/90mmHg more than3 times in 4 times. Otherwise, the blood pressure was unstandard. The blood pressure control of the included population is as follows (see Table 3.2 for details).

## Discuss

“The guideline for the prevention and treatment of type 2 diabetes in China” is a practical guidelines, which were formulated by domestic experts in related fields with reference to the results of the latest large-scale randomized controlled clinical trials and combined with the specific national conditions of our country. The aim of this guideline is to provide evidence-based and effective guidance for clinicians, to ensure that the diagnosis and treatment of diabetic patients get specification, and to avoid the arbitrariness of clinicians in treating patients according to their personal preferences and experience ^[11].^ At the same time, the guideline puts forward the frequency and mode of early screening of prediabetes and diabetes among different population, which can reduce the waste of unnecessary medical resources on the basis of early detection and early intervention. Its ultimate goal is to reduce the prevalence and mortality of diabetes, reduce the incidence of acute complications, and delay the emergence of chronic complications, so as to improve the quality of life of and reduce the disability rate, and then reduce the burden of disease.But according to the overall control of blood glucose in recent years, only formulate a guideline is not enough, the key to the effectively work of the guideline depends more on the clinician’s understanding and mastery of the guideline and whether clinical practice is carried out in accordance with the recommendations of the guidelines. In order to further study the clinical application of the guideline, we investigated the diagnosis and treatment process of all clinicians in a tertiary first-class hospital in Fujian Province. The results showed that there were still serious deficiencies in the standard diagnosis and treatment of diabetes among physicians in various departments. The diagnosis and treatment of diabetes need to be further improved.

### 1. Comprehensive management of blood glucose

Glycosylated hemoglobin is not affected by the occasional fluctuation of blood glucose, and has no significant relationship with the blood sampling time, eating or not, and the use of insulin. It is the gold standard for evaluating the control of blood glucose in the past 2-3 months, also an important basis for guiding clinicians to adjust the antihyperglycemic program ^[12]^. Because of the stability of HbA1c, some people have tried to use it as an index for the diagnosis of diabetes in recent years, and tried to explore the corresponding diagnostic cut-off point in different population ^[13-15]^. In 2010, ADA incorporated glycosylated hemoglobin ≥ 6.5% into the diagnostic criteria of diabetes, which further determined the status of HbA1C monitoring in the diagnosis and evaluation of diabetes. Similarly, China is also constantly exploring the diagnostic cut-off point for diabetes mellitus which were suitable for our country^[16]^. However, due to the determination of glycosylated hemoglobin in China has not yet been standardized, and the diagnostic cut-off point for HbA1C in various regions is not completely clear ^[17]^, it has not been included in the diagnostic criteria of diabetes in China. But it still plays an irreplaceable role in the evaluation of blood glucose control in recent 2-3 months, and in the differential diagnosis of diabetes under stress.

However, in this study, only 60.9% of diabetic patients have monitored HbA1C, in recent 3 months. There are significant differences between different departments in HbA1C monitoring, endocrinology is superior to surgery, internal medicine is superior to surgery. It is showed that the non-endocrine specialist, especially the surgeon didn’t pay enough attention to HbA1C, and not very clear on the importance of glycosylated hemoglobin.They were unable to rational use HbA1C to assess and timely adjustment of antihyperglycemic schemes.

Peripheral blood glucose monitoring can reflect the level of immediate blood glucose and is an effective means to evaluate blood glucose control at and during admission. According to blood glucose fluctuations, clinicians and experienced diabetic patients can independently adjust their diet, exercise and the use of antihyperglycemic drugs, so that blood glucose can be strictly and smoothly controlled in the ideal range and avoid the appearance of hypoglycemia. However, in real life, how many diabetic patients will carry out regular self-blood glucose monitoring?A questionnaire survey was conducted among 2855 patients with type 2 diabetes mellitus. It was found that only 17% of diabetic patients had blood glucose monitoring frequency ≥ 1 time / day, and 31% of patients had blood glucose monitoring frequency ≥ 1 time / week, 14% of the patients had a monitoring frequency of less than 1 time / week, while 38% of the patients never had blood glucose monitoring ^[18]^.In another domestic study, a questionnaire survey among 120 community diabetic patients found that 56.96% of diabetic patients had a blood glucose monitoring frequency of less than 4 times / month ^[19]^.In 2010, Chen Mingdao, Pan Changyu and others conducted a statistical analysis of patients with substandard blood glucose in 181 hospitals among 26 cities across the country. The results showed that among the 3861 people included, only 4.9% of them monitored fasting blood glucose three times a week, 35.8% of the patients were not monitored; only 4.3% of the patients had monitored postprandial blood glucose more than 3 times a week, and 47.8% of them had not been monitored ^[20]^.We can see that most diabetic patients are not aware of the importance of blood glucose monitoring, and very few people have a frequency of blood glucose monitoring more than once a week.While in clinical practice, do we follow the recommendations of the guidelines for blood glucose monitoring? This study showed that regardless of whether the frequency and execution time point of blood glucose monitoring were correct or not, a total of 86.7% (2603) patients had monitored for peripheral blood glucose. Further statistics showed that the average hospitalization days of the blood glucose monitoring group (8.77 ±10.02 days) was significantly less than that of the non-monitoring group (12.79 ±10.25 days),there was significant difference between the two groups (P < 0.05).At the same time, whether or not to monitor blood glucose between departments has statistical significance, endocrinology is better than surgery, internal medicine is better than surgery. This study have shown that there are still about 1/4 clinicians, especially non-endocrine specialists, who are not aware of the role of blood glucose monitoring and are unable to take advantage of it to strengthen blood glucose control, optimize infection control, promote wound healing, shorten the duration of hospitalization and so on.

As we all know, diet therapy is the basic treatment of diabetes. All types of diabetes should adhere to a scientific and reasonable diet, which is a balanced diet, rather than simply controlling food intake, and to match the role of exercise and drugs, and then to achieve good blood glucose, blood lipids and blood pressure control ^[21]^.However, a investigation conducted by Chen mingdao et al showed that among the 3861 diabetic patients whose blood glucose did not reach the standard, only 25.6% insisted on diet control in strict accordance with the doctor’s advice, 58.7% did not follow the strict control, and the remaining 15.7% did not follow the diet control at all ^[20]^.So how many inpatients had received dietary guidance and education after admission?The survey showed that a total of 73.4% (2200 cases) had been given dietary instructions for diabetes, and among the 26.6% (800 cases) who had not received dietary education, 12.5% in internal medicine, 12.5% in surgery, 0.1% in endocrinology and 1.5% in ICU.It suggested that more than a quarter of physicians had not given dietary guidance and education to patients.The average hospitalization days of the two groups were compared according to whether or not dietary education was carried out. It was found that there was no significant difference in the average hospitalization days between the educational group (11.47 ±11.68 days) and the uneducated group (12.55 ±9.74 days) (P > 0.05). This is different from previous studies showing that adherence to dietary therapy can significantly improve blood glucose and glycosylated hemoglobin ^[22]^, and then shorten the length of hospitalization days. Therefore, we should reflect on whether the education content of nurses is in place, whether the patients are effectively implemented, then further investigate the reasons affecting the implementation of patients.

As the disease progresses and the function of pancreatic islet cells declines, it is necessary to consider initiating drug therapy if dietary therapy alone cannot achieve ideal blood glucose control.Because of the wide variety of hypoglycemic drugs and the complexity and diversity of hypoglycemic regimens, we take the relatively simple new type 2 diabetes mellitus as an example to analyze whether the choice of hypoglycemic treatment for hospitalized patients is reasonable.Among the 3000 people included in this study, there were 202 newly diagnosed diabetic patients, 45 of whom met the requirements of HbA1C > 9.0% or FPG > 11.1mmol/L. Among them, only 55.6% (25/45) of the patients received intensive insulin therapy, while 44.4%(20 patients) did not receive intensive therapy, of which 26.7% (12 patients) were in internal medicine and 17.8% (8 patients) were in surgery.It suggested that nearly half of the clinicians could not correctly evaluate the disease status and grasp the timing of insulin initiation therapy.

Surgery group is a special group in hospitalized patients, perioperative blood glucose management is an important indicator to evaluate the implementation of surgeons’ guidelines.Good blood glucose control can promote wound healing and to reduce postoperative infection, poor healing of incision, and the incidence of postoperative cardiovascular complications, reduce patient pain, shorten the hospitalization dates, and reduce the hospitalization expenses. Therefore, clinicians should be familiar with the adjustment of preoperative hypoglycemic scheme, in order to improve the quality of surgery.According to the statistics, a total of 474 inpatients had received major and middle surgery among 3000 patients, among whom 61.8% (293 cases) had been given hypoglycemic treatment before surgery, and only 62.1% (182/293) had been given insulin treatment before surgery.Before the operation, only 41.4% (196/474) of the patients reached the blood glucose standard, and only 51.7% (245/474) of the patients reached the blood glucose standard after the operation.Based on the above analysis, we can concluded that surgeons are weak in perioperative blood glucose management and unable to timely adjust the hypoglycemic scheme, and have a poor grasp of the range of blood glucose control before and after surgery.The purpose of all we have done in diet education, drug therapy and enhanced monitoring is to keep blood glucose stable at the target level, so as to prevent acute complications and reduce and delay the occurrence of long-term complications. How many patients had satisfactory blood glucose control before discharge?Considering the age and complications of the included population, we set a relatively loose range as the boundary. If the peripheral blood glucose meets the requirements of 3.9-10.0mmol/L for ≥3 times in the 4 consecutive monitoring of peripheral blood glucose before discharge, it is defined as good blood glucose control; otherwise, it is considered as poor blood glucose control.The blood glucose monitoring value can be read a total of 2095 patients, of which only 53% (1103/2095) of patients with good standard for controlling blood glucose levels of appeal.Among them, the department of endocrinology is obviously superior to internal medicine and surgery, but there is no significant difference between internal medicine and surgery. The above phenomena suggest that non-endocrine specialists do not have a good grasp of the range of blood glucose standard before discharge, and do not pay enough attention to whether the blood glucose is up to standard at the time of discharge.

### 2. Evaluation, monitoring and prevention of complications of diabetes mellitus

#### 2.1 Control of risk factors for cardiovascular disease

Early studies have found that lipid metabolic disorder is an independent risk factor for cardiovascular disease, which can independently predict the occurrence of cardiovascular events ^[23]^.Due to insufficient insulin secretion and insulin resistance, diabetic patients have a higher incidence of lipid metabolism disorder compared with the normal population.Several studies have shown that strict control of lipids can significantly reduce the risk of cardiovascular events and achieve significant clinical benefits ^[24-26]^.The survey showed that a total of 96.4% (2888cases) of the patients had received blood lipid tests in the past one year. Further statistical analysis of diabetic patients with a clear history of cardiovascular disease showed that of the 714 diabetic patients with cardiovascular disease, only 79.3% (566) were treated with statins. Among the 20.7% (148 cases) patients who did not take statins, 1.4% had abnormal liver function, 2.8% had chronic kidney disease, 7.0% had a history of tumor, and the remaining 9.5% had no obvious statins taboo.Hypertension is not only a common concomitant disease of diabetes, but also a risk factor for cardiovascular and cerebrovascular diseases.Especially when someone combined with the above diseases, the incidence, disability rate and mortality of cardiovascular and cerebrovascular diseases can be significantly reduced by strengthening the control of blood pressure, blood sugar and blood lipids at the same time.The statistics of blood pressure monitoring in the included population were as follows: a total of 2084 diabetic patients with hypertension, among which 89.6% (1868) had blood pressure monitoring;With 140/90mmhg as the boundary, blood pressure reaching the standard was defined as blood pressure below 140/90mmhg for ≥3 times in 4 measurements. The analysis of blood pressure control before discharge showed that there were 1599 patients with complete blood pressure monitoring records, 65.3% (1044/1599) patients with blood pressure meeting the standard at discharge, and 34.7% (555/1599) patients who failed to meet the standard.

Cardiovascular disease is a common macrovascular complication of diabetes, and it is one of the main causes of death in patients with end-stage diabetes.The role of aspirin in preventing recurrence of cardiovascular disease is well recognized.As early as 2009, Baigent C et al. conducted a meta-analysis of 16 secondary prevention studies of cardiovascular disease and found that aspirin reduced the risk of major cardiovascular events by 20% compared with placebo (P<0.01), and did not increase the risk of cerebral hemorrhage ^[27]^.Therefore, many guidelines at home and abroad recommend low-dose aspirin (75 ∼ 162 mg) as a long-term medication for secondary prevention of coronary heart disease.The guidelines also recommend routine use of aspirin as a secondary preventive measure for diabetics with a history of cardiovascular disease.However, in real life, the utilization rate of antiplatelet drugs is not optimistic. A survey on the current situation of aspirin use in four provinces and cities in China shows that the utilization rate of secondary prevention in the total population is 26.61%. The utilization rate of aspirin for secondary prevention of diabetic cardiovascular disease was 51.16% ^[28]^.Further statistics on the population included in our hospital showed that a total of 714 patients had a clear history of cardiovascular disease, of which only 76.6% (548) had received antiplatelet therapy, and 16.1% (115 cases) of the 23.4% (166cases) who had not received antiplatelet therapy had no obvious antiplatelet contraband.It can be seen that the use of aspirin in secondary prevention of diabetes mellitus reported by clinicians in this hospital is higher than that reported in this study, but there is still a gap between them and the guidelines.

#### 2.2 Screening of diabetic complications

Diabetes itself has little effect on the health of patients, but with the progress of the disease, if the blood glucose control is poor, long-term metabolic disorders can cause systemic multi-system, multi-organ complications, which can seriously affect people’s quality of life.Diabetic nephropathy is one of the common and frequent complications, which is the main cause of renal failure in diabetic patients, so early diagnosis and timely intervention is particularly important.Due to the differences in the testing items among the major hospitals, we firstly used the most common index ——serum creatinine, as an evaluation index: in the included 3000 people, nearly 98.1% (2944 people) of patients within 1 year for the monitoring of serum creatinine, only 1.9% of the patients (56) is not monitored.However, it is well known that the human body has two kidneys, and the compensatory function is very powerful, often to severe renal insufficiency, will show the creatinine increases, so the sensitivity is not high.Urinary microalbumin is a more sensitive index for the detection of diabetic nephropathy. Therefore, further statistics have found that only 11.5% (344 people) of the patients have been tested for urinary microalbumin in the past one year. Among them, the department of endocrinology is superior to internal department and surgery, and internal department is obviously superior to surgery.

Diabetic retinopathy is also common microvascular lesions in diabetes, with high specificity, is a common cause of adult blindness, so regular eye assessment is necessary.The results of this survey showed that in the past one year, only 9.6% of the 3000 patients had undergone specialist fundus examination.Among the 90.4% (2713) who did not undergo fundus examination, 51.6% were in internal department, 33.5% in surgery, 2.4% in endocrine department and 2.9% in ICU.

Generally speaking, in our hospital, the monitoring of blood pressure, blood lipid, serum creatinine and blood glucose is more active and standardized, but the screening of diabetic nephropathy and diabetic retinopathy is far from enough.There are still irregularities in the secondary prevention of diabetes. About 1/4 diabetic patients with cardiovascular diseases do not receive antithrombotic treatment, about 1/5 do not receive statins, and about 1/3 patients did not meet their blood pressure when they leave the hospital. The results showed that the clinical implementation of diabetes guidelines is not satisfactory.If the surgical and non-surgical guidelines are defined in accordance with the above evaluation, index monitoring, selection of hypoglycemic regimen, implementation of secondary prevention and perioperative blood glucose management, respectively, the implementation of the guidelines is good, according to statistics, Only 5.6% (168) of non-surgical patients received relatively standardized treatment, and only 0.1% (4) of surgical patients received standard treatment.In a word, the overall implementation rate of the guidelines in our hospital is on the low side, and the mastery and implementation of the guidelines by all clinicians still need to be improved, although this article for single center cross-sectional survey is not enough wide range of aspects, but the sample is representative of the top medical institutions in fujian province, the results of the study have certain reference value.In 2011, Zhou Yingxia and others analyzed the current situation of medical staff at all levels in Shanghai aware of the “guidelines for the Prevention and treatment of Diabetes in China”. It was found that the overall awareness rate of the guidelines was low (37.36%), and the overall correct awareness rate of doctors was higher than that of nurses. The awareness rate of community was lower than that of tertiary and secondary hospitals, and there were significant differences in the mastery of diabetes knowledge among medical staff in different medical institutions ^[29]^.In order to make the guidelines better used in clinical practice, we need to make the following improvements in the promotion and application of the guidelines:1. Increase the holding rate of guidelines, especially in grass-roots hospitals, in order to improve the knowledge level of grass-roots doctors.2. To strengthen the clinical training of doctors, especially non-endocrine specialists, we should increase the knowledge reserve and better guide the clinical diagnosis and treatment by means of professional study, lecture, further study, regular examination and so on.3. Strengthen the professional knowledge training of nursing staff in order to better guide the patients.4. Strengthen the education and behavior guidance of diabetic patients, such as diet education, non-drug treatment, standardizing blood pressure, blood glucose monitoring and so on.5. Implement the follow-up system to ensure the effective implementation of patients. 6. Promote the improvement of the medical system, such as strengthening the reimbursement of medical insurance, improving the two-way referral system and follow-up system ^[30-32]^.

The deficiency of this study: this study is a single-center cross-sectional survey, the source of the sample is relatively limited, it is difficult to represent the implementation of the guidelines of different grades of medical institutions and other hospitals of the same grade. It needs to be further studied by multi-center investigation in the future.Some of the patients are tumor patients, during this period may be admitted to the same medical group, resulting in deviation from the statistical results.Some cadre wards and ICU patients have a long stay in hospital, which has a significant impact on the average length of hospital stay.

## Conclusion

1. Clinicians in this hospital do not pay enough attention to the screening of glycosylated hemoglobin, urinary microalbumin, blood lipids and diabetic retinopathy.

2. Clinicians in this hospital do not have a good grasp of the timing of the initial treatment of insulin.

3. The secondary prevention of diabetic complications by clinicians in this hospital is not in place.

4. The overall implementation rate of diabetes diagnosis and treatment in this hospital is low, and all clinicians, especially non-endocrine specialists, do not have a good grasp of the guidelines.

5. It is necessary to further strengthen the training of clinical medical staff, promote the education of patients’ behavior, promote the improvement of medical insurance, in order to further reduce the incidence of diabetes, reduce mortality, and then reduce the disease burden of diabetes.

## Data Availability

all data included in this study are available upon request by contact with the corresponding author.

